# Novel Missing Data Imputation Approaches Enhance Quantitative Trait Loci Discovery in Multi-Omics Analysis

**DOI:** 10.1101/2023.11.29.23299181

**Authors:** Zining Qi, Alexandre Pelletier, Jason Willwerscheid, Xuewei Cao, Xiao Wen, Carlos Cruchaga, Philip De Jager, TCW Julia, Gao Wang

**Affiliations:** Center for Statistical Genetics, The Gertrude H. Sergievsky Center, Columbia University, New York, NY, USA; Department of Biostatistics, Columbia University, New York, NY, USA; Department of Pharmacology, Physiology & Biophysics, Chobanian & Avedisian School of Medicine, Boston University, MA, USA; Department of Mathematics & Computer Science, Providence College, Providence, Rhode Island, USA; Data Science Institute, Columbia University, New York, NY, USA; Department of Psychiatry, Washington University School of Medicine, St. Louis, MO, USA; NeuroGenomics and Informatics Center, Washington University School of Medicine, St. Louis, MO, USA; Charles F. and Joanne Knight Alzheimer Disease Research Center, Washington University School of Medicine, St. Louis, MO, USA; Department of Neurology, Center for Translational & Computational Neuroimmunology, Columbia University Medical Center, New York, NY, USA; Cell Circuits Program, Broad Institute, Cambridge, MA, USA; Department of Neurology, Columbia University, New York, NY, USA

**Author notes:** Correspondence: Julia TCW,; Gao Wang.

## Abstract

Handling missing values in multi-omics datasets is essential for a broad range of analyses. While several benchmarks for multi-omics data imputation methods have recommended certain approaches for practical applications, these recommendations are not widely adopted in real-world data analyses. Consequently, the practical reliability of these methods remains unclear. Furthermore, no existing benchmark has assessed the impact of missing data and imputation on molecular quantitative trait loci (xQTL) discoveries. To establish the best practice for xQTL analysis amidst missing values in multi-omics data, we have thoroughly benchmarked 16 imputation methods. This includes methods previously recommended and in use in the field, as well as two new approaches we developed by extending existing methods. Our analysis indicates that no established method consistently excels across all benchmarks; some can even result in significant false positives in xQTL analysis. However, our extension to a recent Bayesian matrix factorization method, *FLASH*, exhibits superior performance in multi-omics data imputation across various scenarios. Notably, it is both powerful and well-calibrated for xQTL discovery compared to all the other methods. To support researchers in practically implementing our approach, we have integrated our extension to *FLASH* into the R package flashier, accessible at https://github.com/willwerscheid/flashier. Additionally, we provide a bioinformatics pipeline that implements *FLASH* and other methods compatible with xQTL discovery workflows based on tensorQTL, available at https://cumc.github.io/xqtl-pipeline/code/data_preprocessing/phenotype/phenotype_imputation.html.

## 1. Introduction

Technological advancements in recent years have greatly enhanced the availability of high-throughput biological tools for researchers at reduced cost. This has led to a dramatic increase in the rate of data generation particularly in the area of multi-omics data science, where multiple types of molecular phenotypes (or traits) are generated from the same set of samples. Each type of multi-omics data, such as methylation, proteomics, and metabolomics, offers unique insights into different layers of biological processes. Methylation studies, for example, sheds lights into the epigenetic aspects affecting gene activity. Proteomics focuses on the variety and levels of proteins that emerge from the process of gene expression. Furthermore, metabolomics analyses metabolites as products of synergistic interactions among multiple genes, to provide a broader perspective on metabolic pathways relevant to complex traits and disease phenotypes^[1]^.

Multi-omics phenotypes are now routinely measured in diverse biological samples, spanning various cell types and tissues. These measurements, obtained through different platforms, often contain varying levels of missing data. Such gaps in multi-omics data can occur for multiple reasons: poor sample quality, limited sample size to observe low phenotypic values, technical limitations of measurement platforms, and other reasons for sample drop-out in quality control. Depending on data-type, missing values in multi-omics studies can range from 5% to as high as 80%, according to our literature review detailed in Table S1. Consequently, it warrants thorough investigation how missing data is addressed in multi-omics research. This is particularly crucial given unique nature of each different multi-omics studies that leads to varying degrees and patterns of missing data.

Imputation is a widely accepted strategy for tackling missing data issues, facilitating statistical analyses on complete datasets without excluding samples or features that many statistical methods, not tailored for missing data, cannot handle. Several approaches have been documented in the existing multi-omics literature for missing value imputation^[2]^. Simple methods like mean imputation replace missing values with the average of observed data, whereas the lowest of detection (LOD) method uses the smallest observed value for filling in missing entries. Mean imputation and its more sophisticated derivatives — such as the K Nearest Neighbors (KNN) that leverages closest observed samples — are extensively utilized in various multi-omics studies^[3,4]^. Advanced methods — including low-rank approximation via matrix factorization^[5–7]^ along with sophisticated regression methods that combine single or multiple imputation (SI/MI) with machine learning strategies^[8]^ — have shown better performance than KNN in simulation benchmark studies^[9]^, yet they are not as commonly adopted as KNN or other simpler methods in practice. For a comprehensive list of multi-omics imputation method benchmarks and the recommended approaches derived from these benchmarks, please refer to Table S2.

We have identified several gaps between methodological work in the literature and their practical applications to multi-omics data. Firstly, as indicated in Tables S1 and S2, it is evident that real-world multi-omics studies (even high-impact ones) frequently opt for simpler imputation techniques over more advanced methods^[10,11]^. This trend raises questions about the practical reliability of advanced methods in various multi-omics research contexts, such as clustering, differential expression analysis, and molecular quantitative trait loci (xQTL) discoveries. Secondly, most existing benchmarks focus narrowly on specific types of multi-omics data. There are only a few that comprehensively address a broad spectrum of data types with varying sample sizes, extents, and patterns of missing data^[12–14]^. Furthermore, these benchmarks often rely on synthetic data such as simulations based on real data to assess methods performance, leaving uncertainties about how these findings translate to reliability and robustness in real-world studies. Lastly, to our knowledge there is not yet a benchmark specifically tailored for xQTL analysis. This gap became apparent in our search for methodological guidance to impute multi-omics data for xQTL analysis, posing challenges in making informed decisions for several of our ongoing research projects.

Driven by the practical need to tackle missing data issues in xQTL analysis, we conducted an extensive evaluation of 16 methods as detailed in Table S3 Promising results from some existing methods in our preliminary benchmarks motivated us to develop new enhancements using similar models and algorithms. We utilized data across three cohorts/biobanks, focusing on three types of multi-omics data known for wide-spread missing values (methylation, proteomics, and metabolomics). Our approach involves conducting simulation studies to assess imputation accuracy, xQTL discovery power and calibration, along with real-world xQTL discovery and subsequent replication studies. We show that our extension to a recent empirical Bayes matrix factorization approach, *FLASH*, outperforms other methods in almost every scenario we examined, is reasonably fast for large-scale multi-omics datasets, and producing outputs that are more straightforward to interpret compared to various machine learning approaches.

## 2. Results

### 2.1. Overview of multi-omics imputation and xQTL discovery benchmark design

We developed a comprehensive benchmark for simulation and real data analysis to evaluate methods across three multi-omics modalities — proteomics, metabolomics, and DNA methylation — in three multi-omics resources including the Religious Orders Study/Memory and Aging Project (ROSMAP), Knight Alzheimer Disease Research Center (Knight), and Mount Sinai Brain Bank (MSBB). These datasets underwent standard quality control (QC) and normalization, as previously detailed for these specific datasets^[15–17]^. By offering a diverse range of sample sizes, molecular features, measurement platforms, and missing data patterns, these datasets enables us to create an extensive framework for missing data methods assessment (Details see Section 4).

As illustrated in Figure 1, the multi-omics imputation and QTL analysis benchmark in our study consists of several key steps. We began by simulating missing data patterns completely at random (MCAR) across various rates, from a minimal 5% to an extreme 50%, reflecting both literature reports (Table S1) and the characteristics of our own data-sets (Figure S1). To more accurately approximate real-world multi-omics data with realistic missing patterns, we developed a descriptive model based on data from ROSMAP, Knight, and MSBB to generate missing values not at random (MNAR) for the three modalities assessed (Section 4.2.1). Beyond benchmarking imputation accuracy, we further conducted numerical studies for xQTL discovery. This involved simulating genotypic associations (xQTL) with realistic molecular phenotypes, and applying statistical fine-mapping to determine the impact of imputation methods on identifying true non-zero genotypic effects. We then applied these methods to discover xQTL in our datasets, focusing on pQTL and metaQTL, and compared significant genes identified by each method. To validate the robustness of our findings, we carried out replication studies for pQTL discovery among the three resources.

**Fig. 1.**
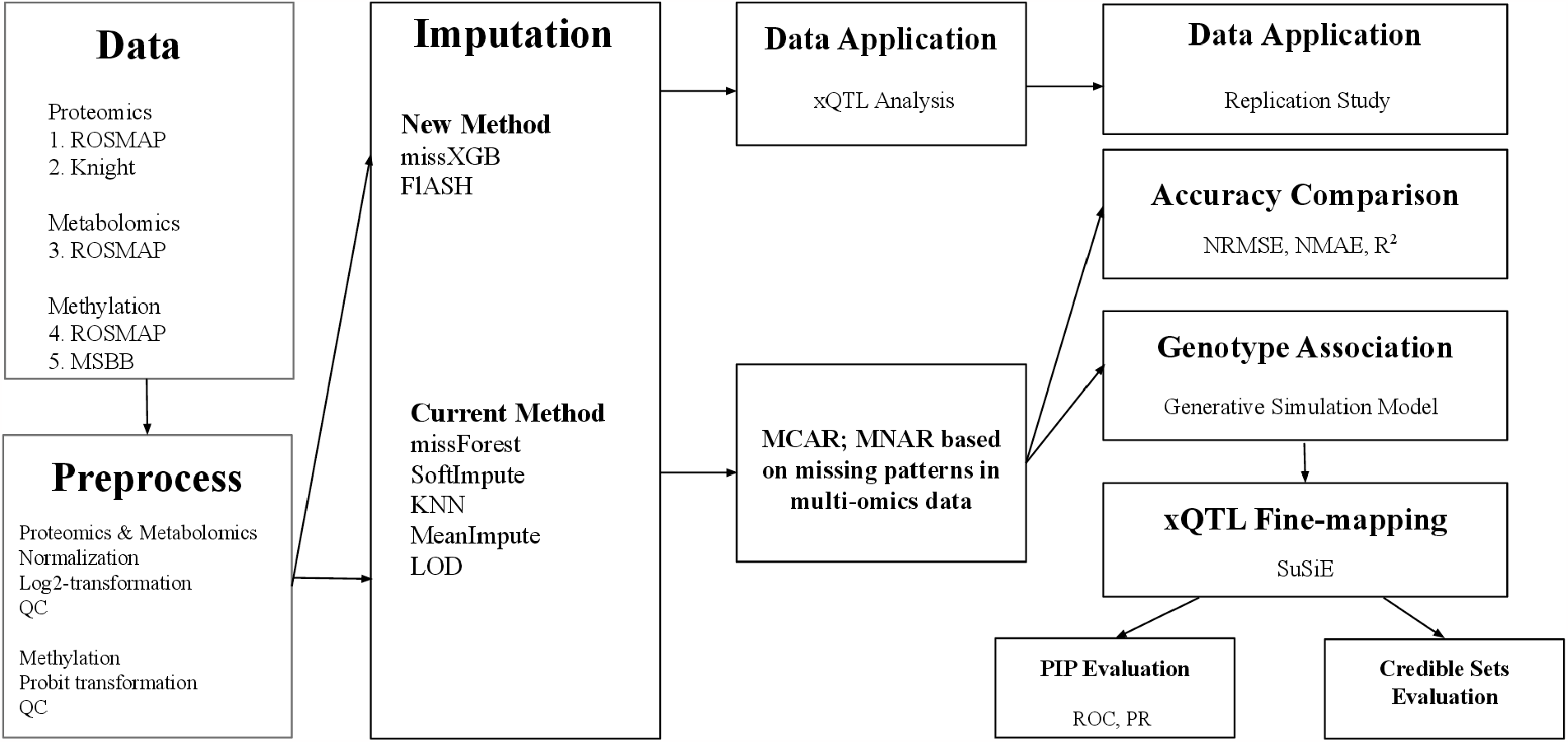
Overview of multi-omics imputation methods and xQTL discovery benchmark. This workflow summarize the datasets, imputation methods and key steps of evaluating imputation methods in the context of xQTL studies, using both simulated and real-world multi-omics data.

We assessed imputation accuracy, xQTL detection power, false discovery rate control, real-world xQTL discovery, replication, and runtime across 16 methods (Table S3). This selection represents major imputation techniques in the multi-omics field (Table S1 and S2), covering: 1) observed value based methods such as *MeanImpute, LOD, KNN* ; 2) low-rank approximation methods such as *SoftImpute* ^[5]^ and *FLASH* ^[18]^; and 3) machine learning regression approaches incorporated to single or multiple imputation frameworks^[19,20]^ such as *MissForest* ^[8]^, and LR^[21]^.

We also introduce two new methods by extending existing ones. The first, *MissXGB*, is an adaptation of *MissForest*, using XGBoost^[22]^ for machine learning regression instead of traditional random forest regression trees. The second, and optimized version of *FLASH*, enhances computational efficiency and imputation accuracy through a genome-wide *FLASH* approach that explicitly models chromosome-specific and global factors. Further details about *MissXGB* and the modified *FLASH* are provided in Section 4.1.

After exploring various parameter settings for the methods under consideration, such as the number of neighbors (*K*) in KNN, the regularization and threshold parameters in *SoftImpute*, and the choice of priors in *FLASH*, we finalized a selection of seven methods for comprehensive benchmarking as described earlier. These methods are summarized with their respective abbreviations in the box “Imputation” on Figure 1. Key findings from our simulation studies are summarized in Figures 2, 3, 4, with additional details in Table S4. Real-world data analysis results are summarized in Figure 5 and Table 1, with additional details in Figures S1 and S2.

**Table 1.** Analysis of pQTL data: discovery from ROSMAP. The first column summarizes the number of pQTL genes identified by each method and their proportion compared to the total gene counts in ROSMAP. The second column compares each method to *KNN*_10_, showing the percentage increase or decrease in the number of significant genes identified. The last column summarizes genes uniquely identified by each method.

| Method | ROSMAP |  |  |
| --- | --- | --- | --- |
|  | #(%) sig. genes | diff. from KNN<sub>10</sub> | method specific |
| *FLASH* | 2,811(37.8%) | 18.8% | 930 |
| *MissXGB* | 2,640(35.5%) | 11.58% | 759 |
| *MissForest* | 2,625(35.3%) | 10.95% | 744 |
| KNN<sub>10</sub> | 2,336(31.4%) | 0.0% | 455 |
| *SoftImpute* | 2,654(35.7%) | 12.17% | 773 |
| *MeanImpute* | 2,585(34.8%) | 9.27% | 704 |
| *LOD* | 2,527(34.0%) | 6.80% | 646 |

**Fig. 2.**
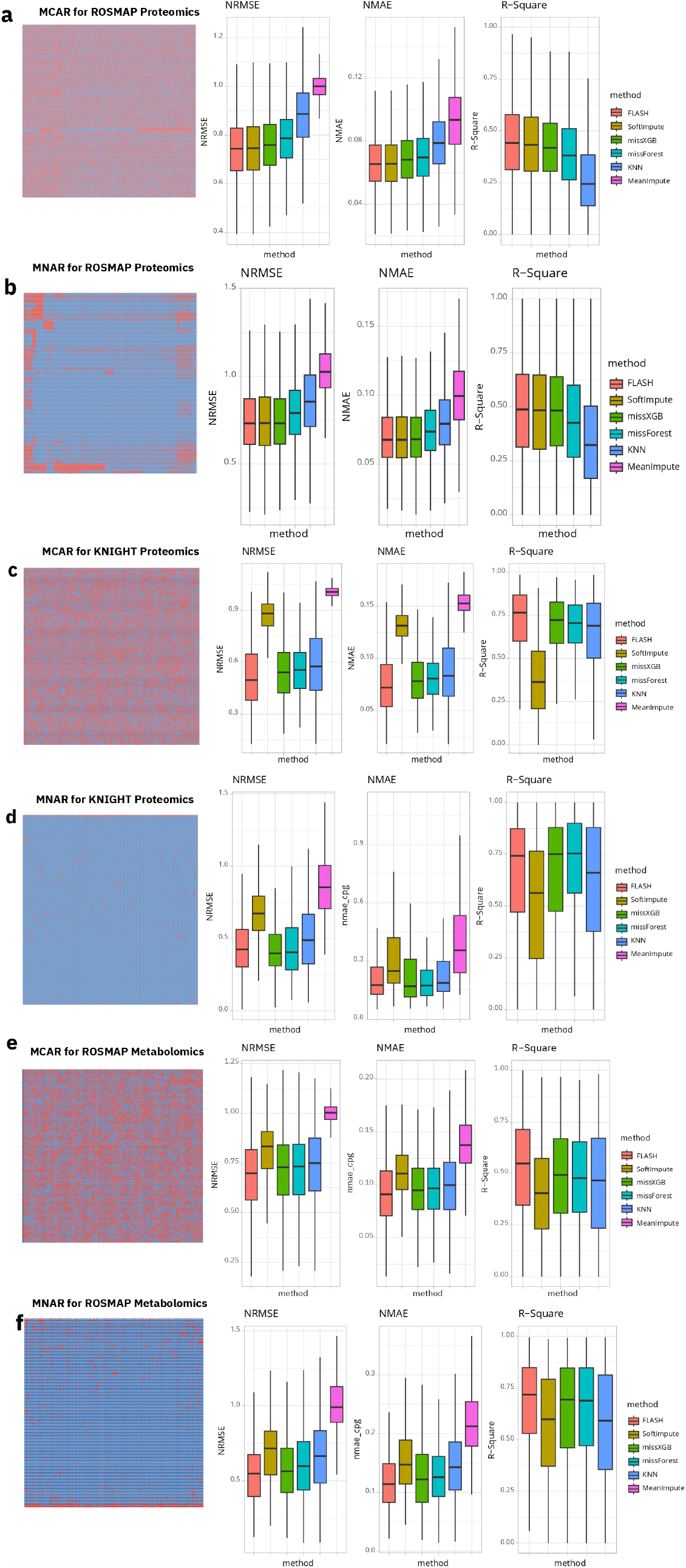
Imputation accuracy for Proteomics and Metabolomics data. Panel a-f summarize the performance of methods on imputation accuracy for proteomics and metabolomics data. Missing patterns are displayed for different scenario and datasets, followed by boxplots showing feature-wise accuracy using 3 metrics. Panel a, c, e illustrate ROSMAP proteomics, Knight proteomics, and ROSMAP metabolomics data with MCAR. Panel b, d, f illustrate ROSMAP proteomics, Knight proteomics, and ROSMAP metabolomics data with MNAR.

**Fig. 3.**
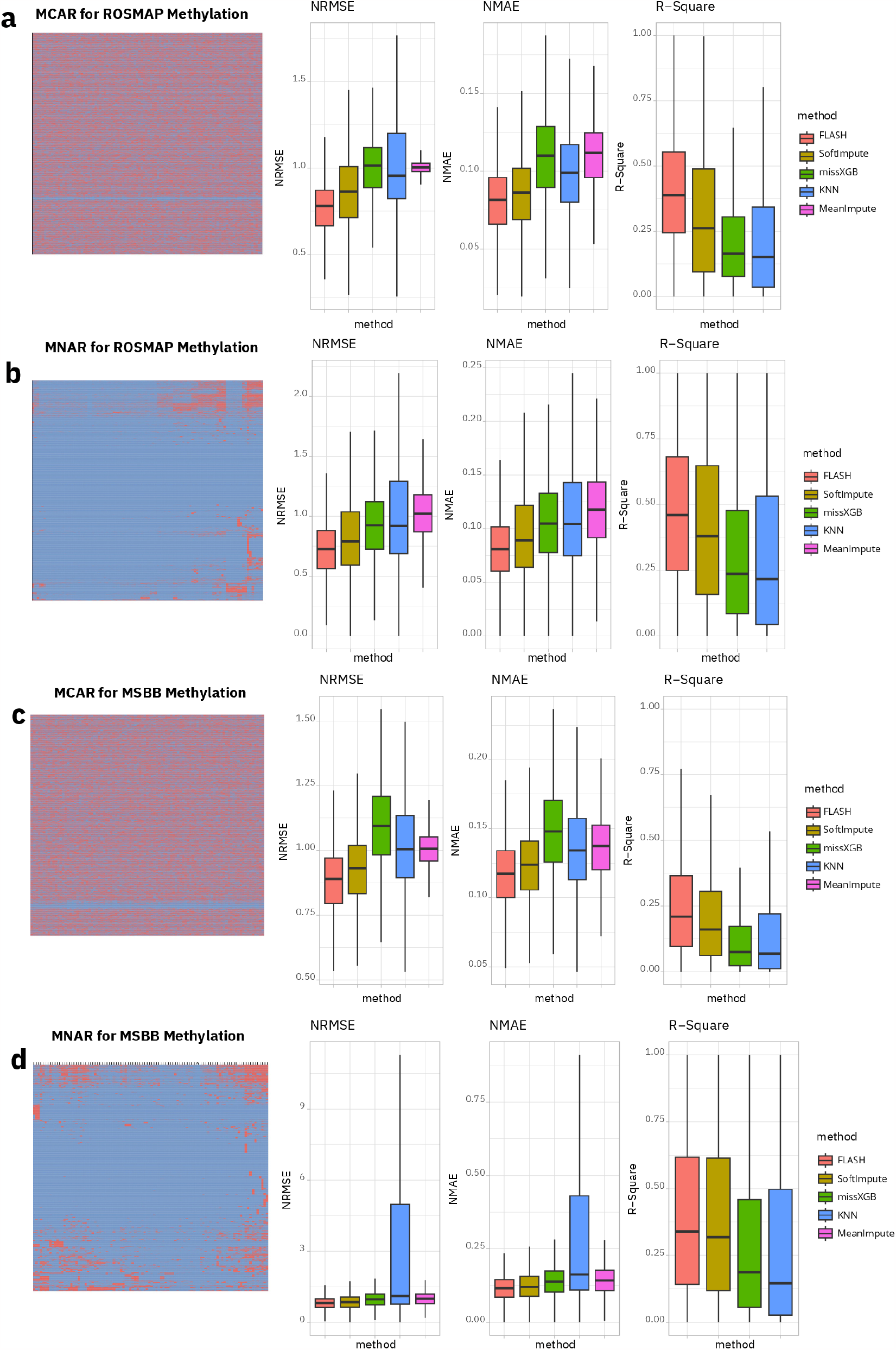
Imputation accuracy for Methylation data. Similar to Figure 2, panel a and c illustrates ROSMAP methylation and MSBB methylation with MCAR. Panel b and d illustrate ROSMAP methylation, MSBB methylation with MNAR.

**Fig. 4.**
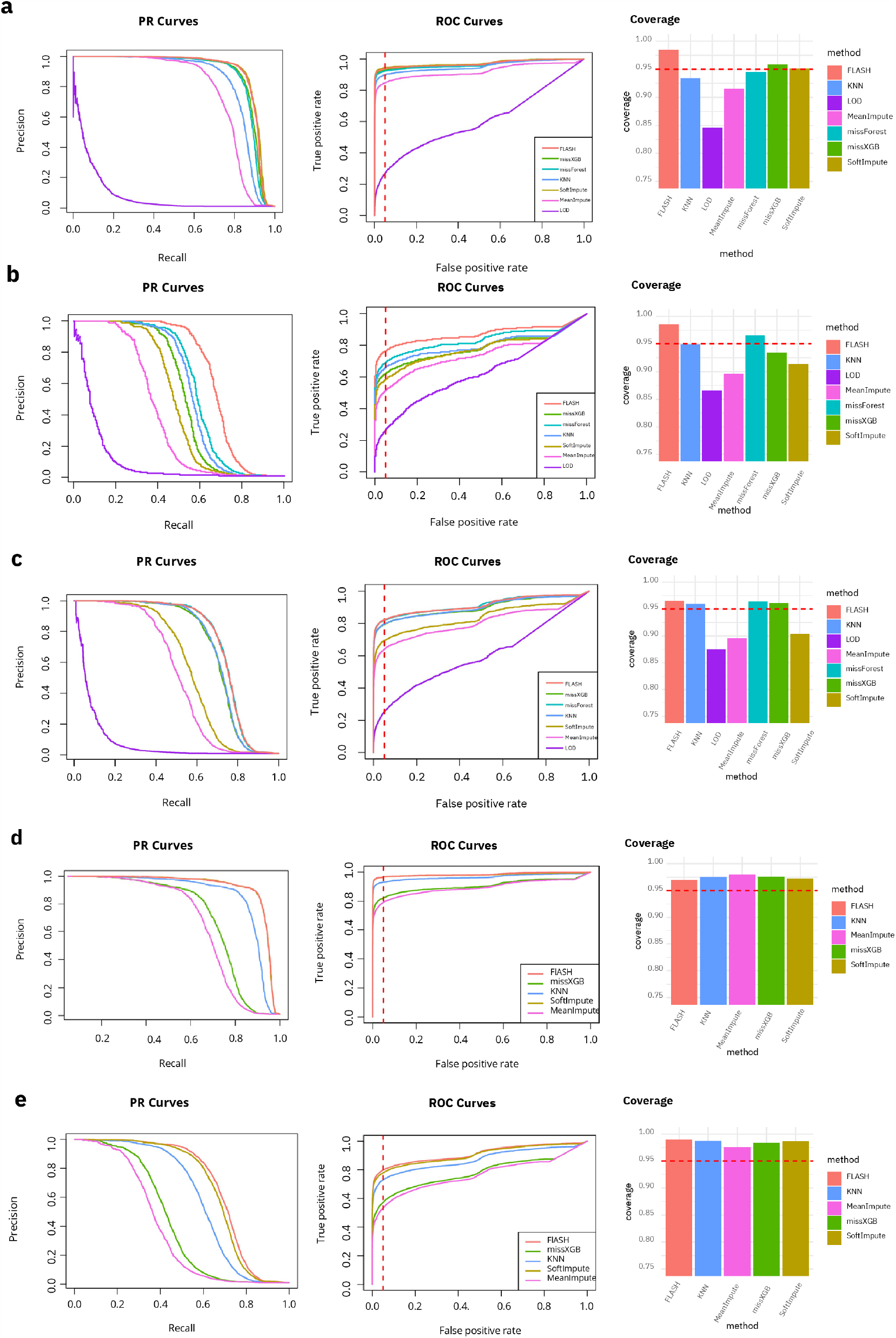
Impact of imputation methods on xQTL discoveries. Fine-mapping posterior inclusion probability and credible sets (CS) comparison of imputation methods are summarized as PR/ROC curves and bar plots. Panel a is results for ROSMAP Proteomics. The PR and ROC curves are calculated based on PIP result from SuSiE^[23]^. The red dashed line on ROC curves are false positive rate at 0.05. Coverage can be interpreted as 1− *FDR*; the red dashed line on the coverage plots is 0.95 corresponding to FDR at 0.05 for fine-mapping CS. Panel b, c, d, e summarize the performance for Knight proteomics, ROSMAP metabolomics, ROSMAP methylation, and MSBB methylation respectively.

**Fig. 5.**
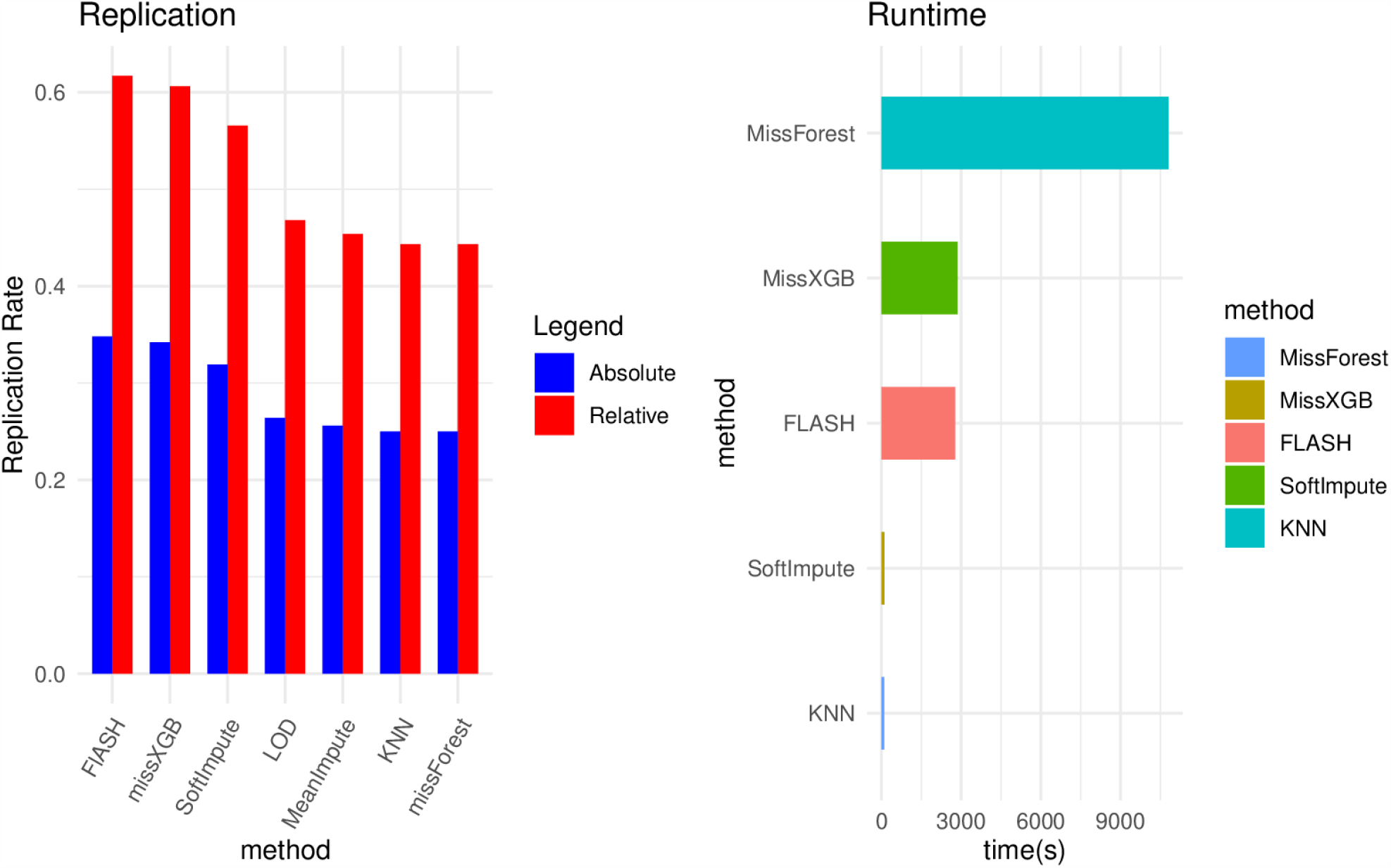
Analysis of pQTL data: replication and runtime. Panel a summarizes the absolute and relative replication rate for each method in the pQTL replication study, comparing discoveries in Knight to those in ROSMAP. The relative replication rate is calculated by absolute replication over replication of joint discovery set. Panel a shows the runtime of each method for imputation on ROSMAP proteomics data.

### 2.2. FLASH outperforms other methods in imputation accuracy for multi-omics data

We gauged the efficacy of various imputation methods using metrics such as NRMSE, NMAE, and *R*^2^, the definitions of which are provided in Section 4.2.2. Our evaluation focused on individual molecular features, including protein levels for specific genes, metabolite measurements, and CpG site intensities. Aggregated performance across multiple molecular features are shown as box plots (Figure 2 and 3). For each dataset we evaluated a range of missing rates of MCAR scenarios, along with dataset-specific MNAR patterns. Interestingly, we observed that the pattern of missing data was less influential than the sample size and the number of features. Therefore, in our analysis, we present MCAR scenarios at 50% as an example of a moderate to high level of missing data, a situation frequently encountered in real-world settings (Table S1, Figures S1 and S2).

We found that widely used observation based methods like KNN (demonstrated here as *KNN* _10_ with *K* = 10) and *MeanImpute* consistently underperform across various scenarios, especially when compared to low-rank approximation and machine learning regression techniques. The new imputation tool we developed, *MissXGB*, showed comparable or slightly superior performance to *MissForest*. We also noted that *SoftImpute* is effective for larger datasets (in terms of samples and features) compared to *MissForest* and *MissXGB*. However, its performance suffers in datasets with fewer samples and features, such as Knight proteomics and ROSMAP metabolomics, though it still surpasses *KNN* _10_ and *MeanImpute*. Our enhanced *FLASH* method proved capable of efficiently handling both large and small multi-omics datasets, consistently outperforming other methods in all evaluated scenarios.

### 2.3. FLASH demonstrates increased power and improved false discovery control in xQTL studies

After conducting fine-mapping on simulated xQTL data, as explained in Section 4.3, we assessed power and false discovery rates using precision-recall and ROC curves at various posterior inclusion probability thresholds. This was done under the challenging conditions of 50% MCAR settings, reflecting moderate to high missing rates commonly seen in practice. Consistent with our predictions, the effectiveness of imputation methods in xQTL discovery power aligns with our earlier evaluation of their imputation performance. For moderately sized multi-omics data, *FLASH* outperforms other methods, closely followed by *SoftImpute*. Both *MissForest* and *MissXGB* show similar results and are notably more effective than *KNN* _10_, *MeanImpute*, and *LOD* (Figures 4, panel a, d, e). Performance of *SoftImpute* declines in smaller data-sets (Figures 4, panel b, c). We noticed that in simulations of Knight-based pQTL studies with small sample and feature sizes, *KNN* _10_ outperforms *MissXGB* in terms of power.

We also evaluated the calibration of fine-mapping credible sets (CS) by their coverage, defined as the proportion of CS capturing the true simulated effects. For well-calibrated 95% CS, the coverage is expected to be at least 95%. Our findings indicate that not all imputation methods lead to well-calibrated CS. As shown in Figure 4, panel a-c, *LOD* and *MeanImpute* have inadequate false discovery rate (FDR) control. *SoftImpute* also fails to control FDR in smaller datasets. Even *KNN* _10_ and *MissXGB* exhibit slightly inflated 95% CS under certain conditions. Reassuringly, both *FLASH* and *MissForest* demonstrate well-controlled FDR. Moreover, compared to *MissForest*, 95% CS from *FLASH* attain higher observed coverage.

The outcomes of our numerical xQTL studies highlight that *FLASH* -based imputation of molecular phenotype measurements is the most powerful for xQTL discovery, while also being the most conservative in terms of FDR.

### 2.4. FLASH yields additional and robust xQTL discoveries in real-world multi-omics data analysis

#### 2.4.1. Statistical analysis for xQTL association

We applied seven imputation methods to proteomics data from ROSMAP (totaling 7,712 genes) and Knight (1,181 genes), along with metabolomics data from ROSMAP (635 metabolites). These datasets were then processed using the FunGen-xQTL analysis protocol described in Section 4.4. Following standard xQTL study practices, we present a gene-level summary of pQTL discovery. This involves permutation testing for each gene within the cis-window to adjust for multiple testing of many genetic variants^[24,25]^, and genome-wide false discovery rate control at gene level using the qvalue method^[26]^. For the metabolomics data, we conducted a GWAS for each metabolite, counting significant metaQTLs as those with p-values below the standard GWAS threshold of 5 *×* 10^−8^, without additional adjustments for multiple testing at metabolites level. xQTL discoveries are summarized in Table 1. We selected *KNN* _10_ as a baseline for comparison due to its popularity in multi-omics literature, and relatively calibrated results across evaluated scenarios as shown in Figure 4.

In the ROSMAP proteomics data, *FLASH* led to the identification of the most pGenes (genes with at least one pQTL), closely followed by *SoftImpute, MissXGB*, and *MissForest*. While *MeanImpute* and *LOD* detected more pGenes than *KNN* _10_, these results warrant caution given their potential for inflated false discoveries as previously demonstrated. In the smaller Knight dataset, *FLASH* proved to be the most conservative, yielding the fewest pGenes. Conversely, *SoftImpute*, which has shown a propensity for false positives, identified nearly double the pGenes compared to *FLASH*, while *MeanImpute* found over three times more. The most striking result was from *LOD*, reporting 1,056 pGenes — 91.7% of all genes in the Knight proteomics dataset — which likely includes many false positives.

#### 2.4.2. Replication analysis for pQTL results between Knight and ROSMAP

In the context of real-world xQTL discoveries, the absence of known ground truth poses a challenge in validating the robustness different imputation methods for the xQTL signals obtained through them. To address this, we conduct replication analyses between Knight and ROSMAP data-sets. Specifically, we designated the pQTL data from the smaller Knight proteomics sample as the discovery set and the pQTL from the larger ROSMAP sample as the replication set. We incorporated cross-method results when defining these sets, aiming to create both reasonable discovery and robust replication data. The criteria for these sets were established as follows:

- Method-specific discovery set: for genes that exist in both ROSMAP and Knight, we evaluate significant genes (permutation q-value *<* 0.05) in Knight pQTL reported by each of the 7 methods, which gives us 7 discovery sets.
- Baseline discovery set: for genes that exist in both ROSMAP and Knight, we consider those reported significant (permutation q-value *<* 0.05) in Knight pQTL by 6 methods excluding *LOD*. This gives us 76 genes.
- Joint discovery set: for genes that exist in both ROSMAP and Knight, we consider those reported significant (permutation q-value *<* 0.05) in Knight pQTL by any of those methods that shows well calibrated coverage in simulation studies, including *FLASH, MissForest, MissXGB* and *KNN* _10_. This is a total of 78 genes.
- Replication set: for genes that exist in both ROSMAP and Knight, we consider those reported significant (permutation q-value *<* 0.05) in ROSMAP pQTL by 6 methods excluding *LOD*. This is a total of 171 genes.
- We compute replicate rate as 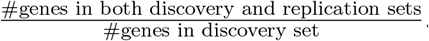. This gives us baseline replication rate of 57.9% and joint replication rate of 56.4%.
- We define relative replication rate as method specific replication rates divided by the joint replication rate.

Method-specific replication rates are shown in Figure 5. In these comparisons, *FLASH* continues to outperform, demonstrating the highest replication rates among the methods evaluated.

Runtime benchmarks for all imputation methods were conducted using the ROSMAP proteomics data. Our findings indicate that while *FLASH* and *MissXGB* take longer to process than simpler methods like *MeanImpute, LOD*, and *KNN* _10_, they are still significantly faster than *MissForest*, and are practical for analyzing real-world datasets.

## 3. Discussion

Missing data in multi-omics studies is a common issue that poses practical challenges. Various algorithms, statistical models, and machine learning methods are theoretically available to impute missing multi-omics data, but beyond simulation studies for imputation accuracy, the impact of missing data and the choice of methods in specific contexts, such as molecular xQTL analysis, is not well-understood. This work demonstrates, through both simulation and data applications, the significant impact of missing data handling on xQTL discoveries. We recommend a Bayesian matrix factorization approach, *FLASH*, as a default choice for imputing missing data in certain types of multi-omics studies including proteomics, metabolomics and methylation.

In our in-depth comparison of the selected seven imputation methods, we evaluated performance across three multi-omics data types with varying sample sizes, number of features, and missing patterns (MAR and MNAR). Our findings align with previously published benchmark on methylation data, which advised against using KNN for imputation and suggested regression-based imputation as a powerful strategy^[27]^. The pattern of missingness appears less critical than the nature of the data (types and size of multi-omics data). For xQTL studies, we found that the performance in power is consistent with accuracy. However, methods like *MeanImpute, LOD*, and sometimes *KNN* _10_, showed inflated type I errors, potentially leading to spurious xQTL signals in practice. In real data analysis, we examined xQTL in multi-omics data from three sources, finding that the choice of imputation method can lead to different genes being identified. In all scenarios and datasets, *FLASH* consistently outperformed other methods in imputation accuracy, power for xQTL discovery, calibration of FDR control, and attained the highest xQTL discovery replication rate. Notably, inferences drawn from *FLASH* were more interpretable than those from other methods, shedding lights into the inherent hidden structures in high-dimensional multi-omics datasets.

The development of new approaches — *MissXGB* and an extension to *FLASH* — was motivated by the need to efficiently analyze large-scale datasets in light of initial promising results from the benchmark. Both *MissForest* and *FLASH*, while more accurate compared to other methods, are slow for large datasets. For example, for the ROSMAP methylation dataset with 721 individuals and 450K CpGs, it takes 7 days for *MissForest* and 2 days for *FLASH* implemented in R package flashier) to complete the analysis. Our new *MissXGB* approach and extension to *FLASH* are much faster while maintaining comparable accuracy. In particular, *FLASH*, as a novel method in the context of multi-omics data imputation, is highly recommended for practical applications.

Our work, while comprehensive, has several limitations that may require further investigation. Firstly, none of the methods or benchmarks we used take advantage of information across different multi-omics modalities, especially when data for the same set of individuals is available, as in the case of the ROSMAP and MSBB datasets in our study with RNA-seq, methylation and proteomics features measured. More sophisticated methods might offer improved results in such scenarios^[13]^, though we did not test their practical performance. Secondly, our analysis was confined to proteomics, metabolomics, and methylation data. We did not assess the impact of missing data and imputation methods on other xQTL data types, particularly single cell or single nucleotide RNA-seq, though there are existing benchmarks and recommendations for these data types^[28]^. Our framework for evaluating xQTL power and FDR could be adapted to these benchmarks to determine whether certain methods can improve xQTL discoveries. Thirdly, the imputation methods we employed are generic. There is potential for future development of methods tailored to specific data types, for example matrix factorization methods — possibly by extending *FLASH* — for functional data such as methylation. We did not explore domain-specific approaches, such as penalized functional regression, which might offer better results for methylation imputation^[29]^. Finally, while our benchmark design is sophisticated and includes both simulation and real-data applications, the methods we considered are limited to those that are popular and computationally feasible. In particular, we did not systematically evaluate deep learning approaches, which might be effective when executed properly^[30,31]^. In fact, we did implement and benchmark a variational auto-encoder approach^[32]^, but found its performance to be highly sensitive to parameter tuning and generally less satisfactory than simpler methods, leading us to exclude it from our in-depth comparison.

Implementation of our benchmark outlined in Figure 1 is available at https://github.com/zq2209/omics-imputation-paper. The R package *MissXGB* is available at https://github.com/zq2209/missXGB. Our extension to *FLASH* is available at https://github.com/willwerscheid/flashier. Imputation pipeline for real-world data analysis implementing seven methods can be found at https://cumc.github.io/xqtl-pipeline/code/data_preprocessing/phenotype/phenotype_imputation.html.

## 4. Material and Methods

### 4.1. Multi-omics missing data imputation methods

In this manuscript, we focus on several key methods, including *MeanImpute, LOD, KNN* _10_, *SoftIm-pute*, and *MissForest*, each with different parameterizations as detailed in Table S3. Additionally, we introduce two new approaches, *MissXGB* and *FLASH*, detailed in the subsequent sections.

#### 4.1.1. The MissXGB algorithm

We developed the *MissXGB* algorithm and software package in R, which follows from the *MissForest* ^[8]^ framework but using an XGBoost model trained on the observed portions of a dataset to predict missing values. Specifically, a data matrix *D*_*n×p*_ can be divided into four parts based on any given *j*-th variable *D*_*j*_ (a length-*n* vector): *Y*_*obs*_, *Y*_*miss*_, *X*_*obs*_, and *X*_*miss*_. Here, *Y*_*obs*_ represents the observed values of *D*_*j*_, while *Y*_*miss*_ denotes its unobserved values. Variables other than *j* in the dataset are partitioned into *X*_*obs*_ amd *X*_*miss*_ for observed and missing values respectively. Overall, for each column *X*_*miss*_, a XGBoost model is trained with response *Y*_*obs*_ and predictors *X*_*obs*_. The trained model is then applied to predict *Y*_*miss*_ with *X*_*miss*_. The iteration will stop as soon as the difference between the newly imputed data matrix and the previous one increases for the first time. Algorithm 1 outlines the *MissXGB* implementation.

##### Algorithm 1

*MissXGB* Algorithm

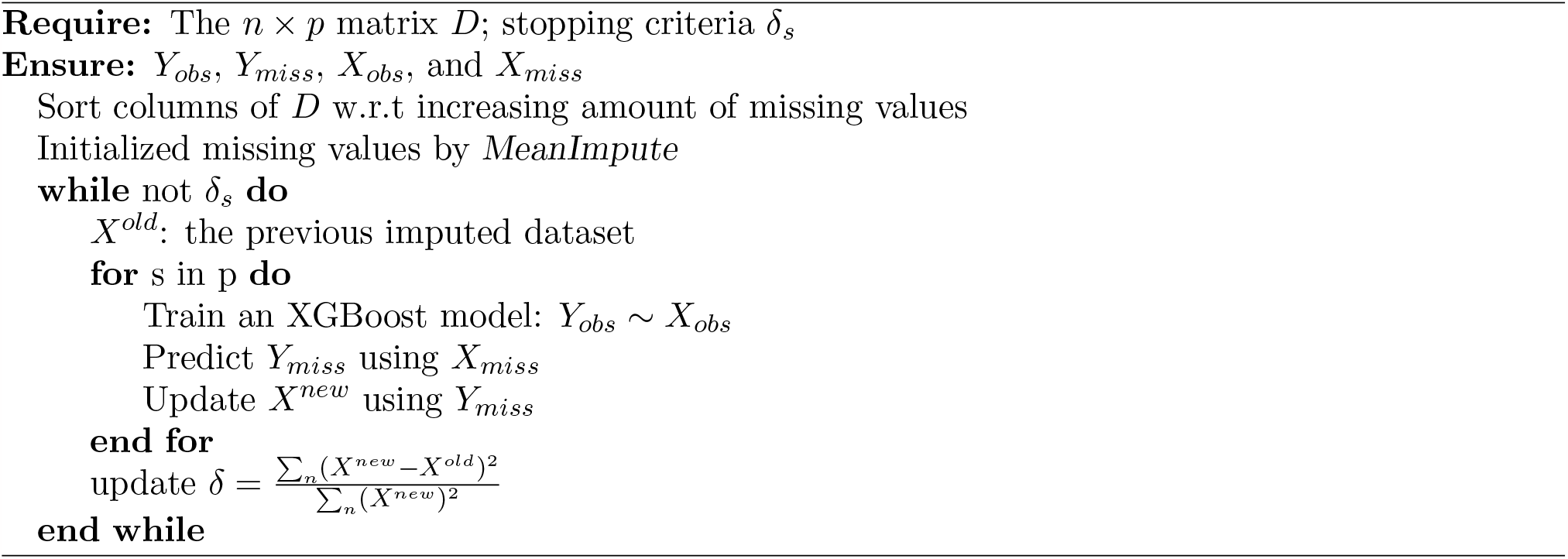

#### 4.1.2. Extension to FLASH

*FLASH* — the Factors and Loadings by Adaptive SHrinkage — is an Empirical Bayes Matrix Factorization (EBMF) model that can be applied to perform low-rank approximation for high-dimensional data. We apply *FLASH* to identify hidden structures in datasets and use them to impute missing values. Unlike standard matrix factorization methods like PCA or SVD, *FLASH* offers a more versatile Empirical Bayes approach. This flexibility allows the model to handle various levels of sparsity in the loadings (*L*) and factors (*F*). One advantages is its adaptive shrinkage feature, which automatically selects the number of factors for *L* and *F*, saving users from having to manually choose and adjust this parameter.

### 4.2. Numerical Study on imputation accuracy

#### 4.2.1. Simulating realistic missing patterns in multi-omics data

In our study, we assessed the performance of different methods using simulated datasets with missing values categorized as either completely at random (MCAR) or missing not at random (MNAR)^[33–35]^. The MNAR simulations were crafted to mirror missing patterns found in actual multi-omics data. We implemented a Bayesian approach to create these missing values, taking into account both the observed data and the identified missing patterns. By clustering samples and features based on their missing profiles, we identified major patterns of missingness in data. The probability of occurrence of missing patterns was used as a prior. This, in conjunction with the observed probability of data being missing, enabled us calculate the probability of missingness taking into consideration of information both from other variables (common in missing at random, MAR, scenarios) and from the variable itself (a characteristic of MNAR situations).

To implement this, consider data matrix *D* with dimensions *n× p*. This matrix includes *n* observations and *p* features, and contains the missing data patterns we aim to capture using simulated data. To create generative models from these patterns, we characterize them using two steps of clustering. First, we group the *p* features into *g* clusters using a *K*-means algorithm. For instance, in methylation data, we use *g* = 100. This gives us a new matrix of size *n× g*, showing the average missingness for each cluster of features. Next, we cluster this matrix by samples using hierarchical clustering with a cutree algorithm^[36]^, forming *k* sample clusters. In the case of methylation data, we set *k* = 10. This creates a total of *g × k* unique clusters, each representing a different missing data pattern.

To apply the derived missing pattern cluster to a new dataset *D*^*′*^, we first assign each individual in *D*^*′*^ to one of the *k* sample clusters based on the relative frequency of occurrence of the sample cluster in dataset *D*. Then, for each of the *g* feature clusters, we compute the probability of a data point from cluster (*k, g*) being missing as

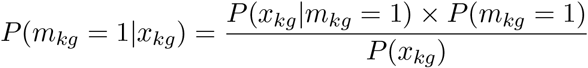

where *P* (*m*_*kg*_ = 1) is the probability of missing data in cluster (*k, g*), estimated by the proportion of missingness in this cluster. *P* (*x*_*kg*_) is the probability density of values in cluster (*k, g*), which can be approximated from the observed data. *P* (*x*_*kg*_ |*m*_*kg*_ = 1) is the probability density of missing values, approximated using features with higher missing rates within the cluster, defined as those having missing rates above the median for features with at least one missing value. To compute *P* (*x*_*kg*_) and *P* (*x*_*kg*_| *m*_*kg*_ = 1), we consider the area under the curve (AUC) of the empirical density function, within one standard deviation around the mean values of these variables, i.e., AUC within the range of 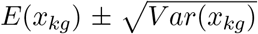 and 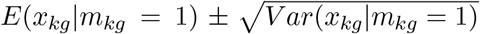, respectively. This process is iteratively applied to each feature within every cluster, until a sample is simulated to accurately reflects the intended missing pattern for the features. Other samples in *D*^*′*^ can be simulated similarly.

#### 4.2.2. Imputation efficacy metrics

In our analysis, we evaluated the effectiveness of various imputation methods using three key metrics: normalized root mean square error (NRMSE)^[37]^, normalized mean absolute error (NMAE)^[38]^, and the squared Pearson correlation coefficient (*R*^2^) that we computed at feature level. NRMSE, defined as the root mean square error normalized by the standard deviation of the observed value of the feature, is calculated as 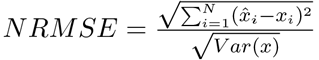 where *N* is the number of observations with missing value, *x* is the observed values, and 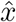 is the predicted values. NMAE, the mean absolute error normalized by the range of the observed value, is given by 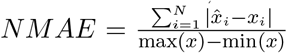. The *R*^2^ metric measures the linear correlation between observed and predicted values, represented by the squared Pearson correlation, defined as 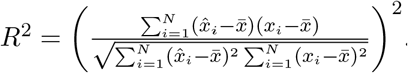

### 4.3. Numerical study on xQTL discovery

#### 4.3.1. Simulate genotype and molecular phenotype association

To accurately capture the molecular phenotype structure found in real multi-omics data, we devised a reverse simulation approach generating genotype of variants in cis-regulatory regions in linkage disequilibrium (LD) with each other, and their association with molecular features observed in multiomics resources previously described. We considered five phenotype datasets, listed in “Data” of Figure 1, to conduct the simulation study. Following basic processing procedures (elaborated in Figure 1), we select subsets of molecular features for proteomics and metabolomics data requiring that 1) features have no missing entry for MCAR based simulation scenarios; 2) features with less than 5% missing rate in the MNAR scenario. For methylation data containing many CpG sites, we compute PCA of CpGs in each topologically associating domain (TAD)^[39]^ and select up to 10 CpG most correlated to the first PC with *R*^2^ *>* 0.75.

For each molecular feature (a gene, a metabolite or a CpG site), we considered the generative model *y* = *Xb* + *Zc* + *ϵ*, where *X* is an *N × P* genotype matrix for *P* variants, *b* is a *P ×* 1 vector of the effect size on molecular genotype on *y*, and *ϵ* ∼ *N* (0, *σ*^2^) is other random effects. After regressing out known and inferred covariates *Z* from *y* ^[40]^ we generate genotype by using this residual *y*_*res*_ ^[41]^. Specifically,

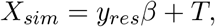

where *T* ∼ *MV N*_*p*_(0, Σ) is a multivariate normal distribution with mean 0 and covariance matrix Σ being the LD matrix of genotype. For each molecular phenotype, we randomly drew *M*_*block*_ ∈ [5, 15] LD blocks and *P*_*m*_ ∈ [10, 50] variants in each LD block for *m* = 1, · · ·, *M*_*block*_. Note that 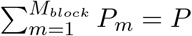. For *m*-th LD block, we used the following procedure to generate LD structure:

1. Generate a Barabasi-Albert network (BAN) to mimic LD among variants^[42]^. If two variants are connected in the BAN, these two variants are considered to be in LD; otherwise, these two variants are considered independent. The variants may be connected only if they are in the same LD block.
2. Let Θ = (*θ*_*pk*_) be an initial concentration matrix for *p, k* = 1, …, *P*_*m*_. Here, *θ*_*pk*_ is set as 0 if two variants (*p, k*) are not connected in BAN; *θ*_*pk*_ from a uniform distribution on the domain of [−0.9, −0.1] ∪ [0.1, 0.9] if two variants (*p, k*) are connected in BAN.
3. Rescale the non-zero elements in Θ to assure its positive definiteness, that is, we divide each off-diagonal element by *λ*-fold of the sum of the corresponding row, where *λ >* 1.5 is the rescale rate. Then, the rescaled matrix is averaged by its transpose to ensure symmetry.
4. Denote *W* = (*ω*_*pk*_) as the inverse of Θ after rescaling and averaging. Elements Σ_*pk*_ in the covariance matrix Σ is determined by 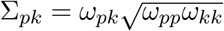.

We randomly selected one to five linkage disequilibrium (LD) blocks from *M*_*block*_ to establish the cis-windows for xQTL analysis. Within each block, one variant is randomly chosen as the true causal variant, with a fixed effect size *β* = 1. The residual variance *σ*^2^ is used to adjust the proportion of variance explained (PVE) by genetic variants for a molecular phenotype. By definition, for a generative model *y*_*sim*_ = *y*_*res*_ + *ϵ* where 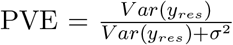. In our reverse simulation model, 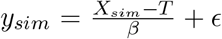, leading to 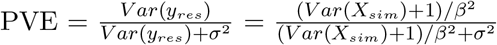. For proteomics and metabolomics in our simulation model, we assume a total PVE of 25% (per variant PVE is 5%), while for the methylation study, a 50% PVE^[43,44]^ is assumed (per variant PVE is 10%). This approach allows us to simulate a realistic scenario reflecting the genetic contributions to molecular phenotypes in these studies.

We conducted multiple independent simulations to evaluate power and false discovery rate (FDR). Each molecular feature in the simulation is treated as an independent analysis unit. For the ROSMAP proteomics data, we considered 3,851 features across 3 replicates, which amounts to 11,553 association analysis units for assessing power and FDR. The Knight proteomics dataset, having a much fewer number of features 49, required 10 replicates to ensure a robust evaluation of power and FDR for this particular dataset with a total of 490 analysis units. The ROSMAP metabolomics dataset has 369 features across 5 replicates, amounting to 1845 associations analysis units. We analyzed 1,381 TADs for methylation data, with each TAD containing 10 CpG sites. This results in a total of 13,810 association analysis units to robustly evaluate methylation QTL. We combine these molecular features into matrices *Y* = [*y*_1_, *y*_2_, …, *y*_*R*_] and assign missing data to them based on MCAR model with 50% missing rate, to assess a moderate to high missing data scenario.

#### 4.3.2. Statistical analysis for xQTL association

Statistical fine-mapping of the simulated genotype across the three molecular phenotype types was conducted on each replicate using SuSiE^[23]^. Fine-mapping serves to address multiple testing issues in cis-xQTL associations, and to differentiate causal variants from other variants in LD with them. The outputs of fine-mapping are posterior inclusion probabilities (PIPs) and credible sets each capturing a single causal variants. To evaluate these results, we generated precision-recall (PR) and ROC curves using the R package ROCR^[45]^ at various PIP thresholds. Additionally, we assessed the coverage of credible sets to evaluate the false discovery rate for single effects fine-mapped. Further details on these evaluation methods are available in the SuSiE manuscript^[23]^.

## Data Availability

All data produced in the present study are available upon reasonable request to the authors

## Supplementary Figures

We validate our realistic missing generation approach on real data. The following is the heatmap of the observed and generated missing patterns on the data.

**Fig. S1.**
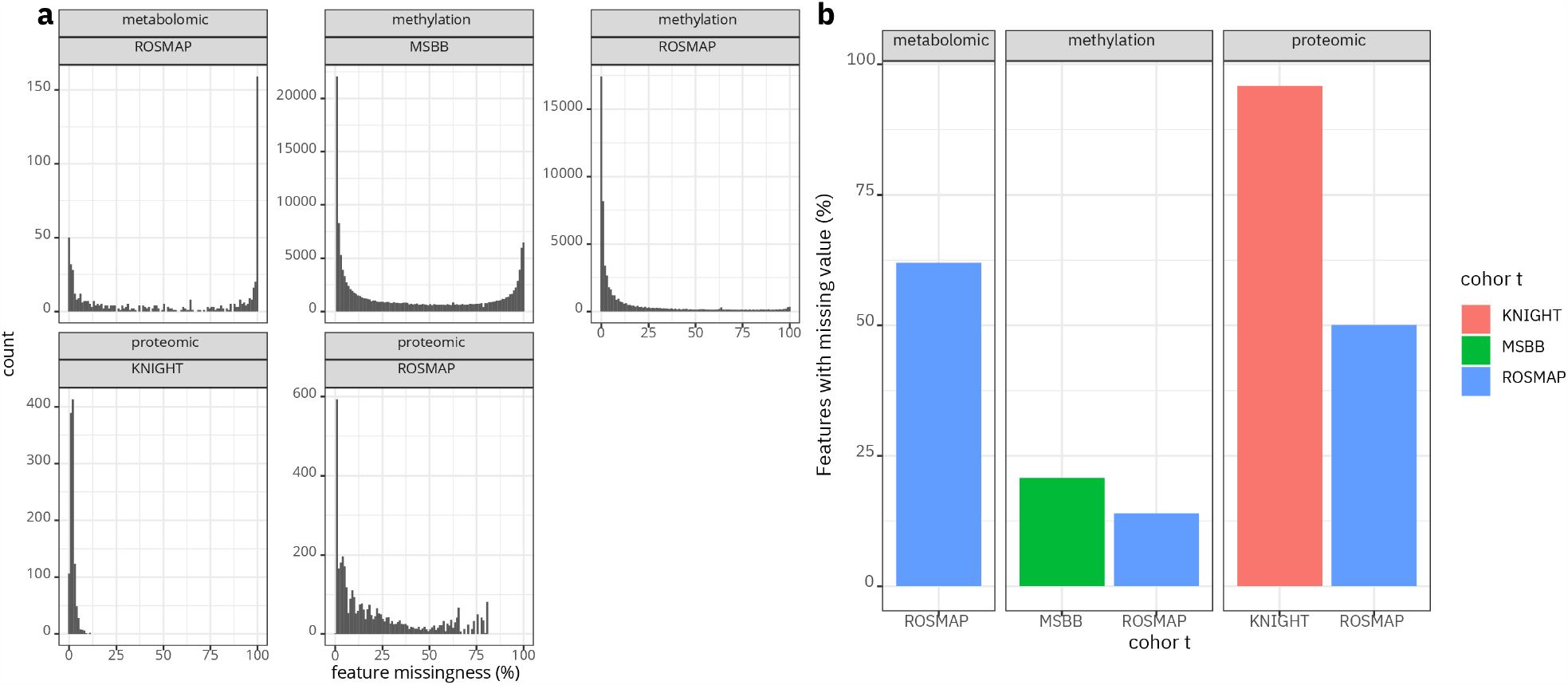
missing rate distribution. Panel a-b summarize the missing rate distribution for datasets. Pabel a is the summary of distribution of missing rate for features across datasets. Panel b summarize the proportion of features that have at least one missing entry across datasets.

**Fig. S2.**
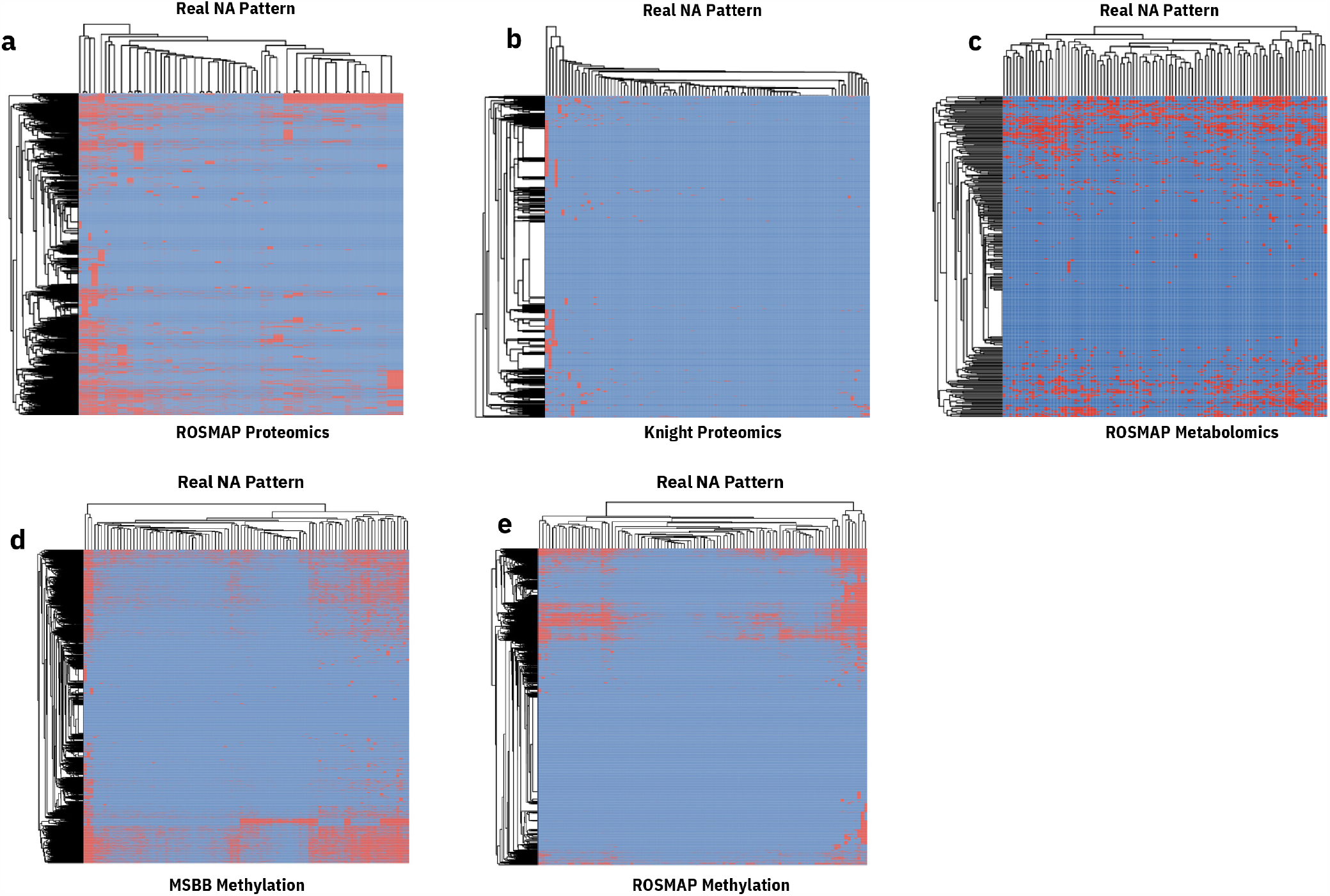
Real Missing Pattern. The heatmap summarize the onserved missing for each datasets. Panel a is the observed missing pattern for ROSMAP proteomics, panal b for Knight proteomics. Panel c summarize the missing pattern for ROSMAP metabolomics. And panel d-e are missing pattern for MSBB and ROSMAP methylation.

## Supplementary Tables

**Supplementary Table S1**: Multi-omics Application Literature Involving Imputation Methods

**Supplementary Table S2**: Methodology and Benchmarking Literature for Imputation Methods

**Supplementary Table S3**: List of Imputation Methods Evaluated

**Supplementary Table S4**: Sensitivity of Tuning Parameter Settings in Imputation Methods

